# Physiological and Perceptual Responses Across Menstrual Cycle Phases in Female Ultra-Endurance Athletes: A Pilot Study

**DOI:** 10.1101/2025.11.13.25340155

**Authors:** I. Molina-Gonzalez, S. R. Gray

## Abstract

Ultra-endurance places considerable physiological stress on the body, and understanding how hormonal fluctuations affect women, who take part in ultra endurance training and events, could improve both safety and performance. The aim of this pilot study was to explore physiological and perceptual responses across MC phases in trained female ultra-endurance athletes.

**Methods:** Naturally menstruating participants (VO₂max: 51.4 ± 5.7 mL·kg⁻¹·min⁻¹; ultramarathon experience: 2.6 ± 1.1 years) were recruited. Seven completed a questionnaire on past experiences and performance perceptions related to the MC. Six underwent physiological testing in the early follicular (EFP), late follicular (LFP), and mid-luteal (MLP) phases. Assessments included resting metabolic rate (RMR), heart rate variability (HRV), and an incremental treadmill test for submaximal and maximal performance. Phase-specific questionnaires recorded perceived performance and well-being.

**Results:** More than half of participants (57.1%) reported perceived performance decrements in the days preceding and during menstruation. However, no significant differences were found in submaximal (heart rate, running economy, rate of perceived exertion) or maximal (VO₂max, peak velocity, time to exhaustion) physiological markers across MC phases. Respiratory frequency at LT1 showed a significant main effect of phase (p = 0.045), though pairwise comparisons did not reach significance. Resting variables, including HRV and RMR, or well-being markers, did not vary significantly between phases.

**Conclusions:** Physiological performance remained stable across MC phases, though trends suggested improved ventilatory efficiency in the LFP compared to the MLP during submaximal work. Despite the small sample size, the frequent reporting of MC-related symptoms indicates the importance of personalised MC monitoring. These findings highlight the practicability of integrating both objective and perceptual measurements in ultra-endurance research and support further investigation with larger cohorts.

## 1. Introduction

Women have historically been underrepresented in sport and exercise science and medicine research, and this gap continues to limit the validity and applicability of current evidence. Foundational physiological studies have been largely conducted on male participants, resulting in training, nutrition, and performance guidelines that are primarily based on male data. Although female inclusion has modestly increased in the last decade, from 39% to 44%, only 5.6% consider the menstrual cycle as a biological factor in study design^1^. This is despite it being known that hormonal fluctuations which occur in the MC influence metabolism^2^, cardiovascular and autonomic regulation^3,4^, thermoregulation^5^, substrate metabolism^6^ and recovery capacity^7^. This gap in sex-specific evidence raises concerns about the applicability of male-derived data to female athletes, particularly within endurance disciplines that impose significant physiological stress.

Ultramarathons are events that exceed the standard marathon distance of 42.2 km (26.2 miles) and impose additional physiological demands that differ substantially from those observed in shorter endurance events. Ultramarathons involve sustained submaximal effort lasting from several hours to multiple days, leading to pronounced metabolic, cardiac, hormonal, thermoregulatory and neuromuscular strain^8–11^. For example, during a 24-h ultramarathon, female runners can experience energy expenditures exceeding ∼45MJ (∼10755Kcal)^8^ and body mass losses of 2.4-4.4%^8^. Despite growing female participation in such events, e.g. rising from 25% in 2022 to nearly 29% in 2024 at the Ultra-Trail du Mont-Blanc (UTMB)^12^, female-specific evidence remains limited. Understanding how hormonal fluctuations across the MC influence physiological responses under these extreme conditions is therefore essential to inform evidence-based strategies for training, recovery and performance optimisation.

The typical eumenorrheic MC lasts 21 to 35 days and includes two main phases, the follicular and luteal, separated by ovulation^13^. Oestrogen dominates in the late follicular phase (LPF), whereas higher levels of progesterone are observed during the mid-luteal phase (MLP), both hormones regulating systemic physiological effects^13^. Oestrogen enhances mitochondrial efficiency, increases lipid oxidation, glycogen storage, and endothelial function^14–17^, whereas progesterone may exert opposing, catabolic effects, increasing thermogenesis, cardiovascular and ventilatory responses^18,19^. The crosstalk between these hormones affects substrate utilisation, fatigue resistance, and recovery kinetics – factors critical to endurance performance. Indeed, oestrogen’s protective effects against exercise-induced muscle damage^20,21^ and its modulation of nitric-oxide-mediated vasodilation^16^ may provide phase-dependent performance advantages. However, findings from existing literature remain inconsistent: some studies report enhanced fat oxidation and glycogen sparing during high-oestrogen phases^6^, while others have observed lower lactate production and improved clearance in the luteal phase, which may explain reports of better performance in marathon distances during this phase^22–24^. Conversely, several studies have found no significant phase-related difference in substrate utilisation or overall performance^13,25^

Conflicting findings on the effects of MC on performance largely reflect methodological inconsistencies: small sample sizes, inconsistent phase identification, and variable study designs with different durations and exercise intensities. Schaumberg et al. (2017) proposed a three-step verification method (calendar tracking, urinary LH testing, and serum hormone), now considered the gold standard, though its use remains limited by practical constraints^26^. Studies adopting this approach, such as the FENDURA Project, found no group-level differences in VO_2_peak, running economy, or power output across MC phases in endurance-trained female athletes, but presented substantial inter-individual variability, suggesting the need for personalised approaches^27^. However, these data derive from endurance, not ultra-endurance, athletes—whose chronic high-volume training, energy deficits, and prolonged submaximal work may interact differently with hormonal fluctuations.

While objective performance markers often appear stable, perceptual factors frequently vary. Menstrual symptoms—including fatigue, pain, and mood changes—are commonly reported as barriers to optimal training and competition^28^. In a six-month longitudinal study, 40% of elite British track-and-field athletes reported reduced performance during the MLP and 35% during the EFL^29^. Such findings highlight that psychological and perceptual dimensions are essential when supporting female athletes.

Understanding the effect of the MC on performance remains to be elucidated. Ignoring its physiological and perceptual effects risks perpetuating male-training models and compromising female health and potential. Moreover, most of the MC-related studies are in endurance models, and this limits the applicability to ultra-endurance contexts, where the duration, intensity and cumulative training stress differ substantially^8–11^. These unique demands may influence how hormonal fluctuations across the MC affect performance and recovery, yet this has never been explored.

The aim of this pilot study, therefore, was to explore physiological and perceptual responses across MC phases in trained female ultra-endurance athletes.

## 2. Materials and Methods

### Participants

Ten healthy, eumenorrheic (mean menstrual cycle length: 28 ± 7 days) female ultra-endurance athletes, aged between 18 and 45 years, volunteered for this investigation. They were recruited through running clubs, social media platforms, and word of mouth. Eligibility criteria included a minimum of two years of ultramarathon experience, ongoing weekly training exceeding 7 hours, and no use of hormonal contraceptives for at least three months preceding the study. Participants were excluded if they reported irregular cycles, menstrual dysfunction such as amenorrhoea or polycystic ovarian syndrome, perimenopausal symptoms, pregnancy, or any metabolic or cardiovascular disorder that might influence testing outcomes. Of the ten athletes enrolled, six completed the full testing protocol, and seven responded to the performance perception and past-experiences questionnaire.

### Experimental Design

A repeated-measures design was applied, consisting of four sessions. During the initial session, participants were familiarised with all study protocols, completed informed consent, underwent health screening via the PAR-Q+, and provided anthropometric data. They also filled in a questionnaire assessing MC–related perceptions of performance and previous experiences (Supplementary Material).

Testing sessions were performed during different menstrual cycle (MC) phases: EFP, LFP, and MLP. All tests were conducted within one to three consecutive cycles. During each session, resting metabolic rate (RMR), heart rate variability (HRV), and performance during an incremental treadmill test were measured.

The MC phase was identified by combining calendar-based tracking and urinary ovulation testing. Athletes were instructed to maintain their usual training and competition schedules throughout the study.

Ethical approval was granted by the University of Glasgow College of Medical, Veterinary and Life Sciences Research Ethics Committee for Non-Clinical Research Involving Human Participants (Ref: 200240251).

**Figure 1:**
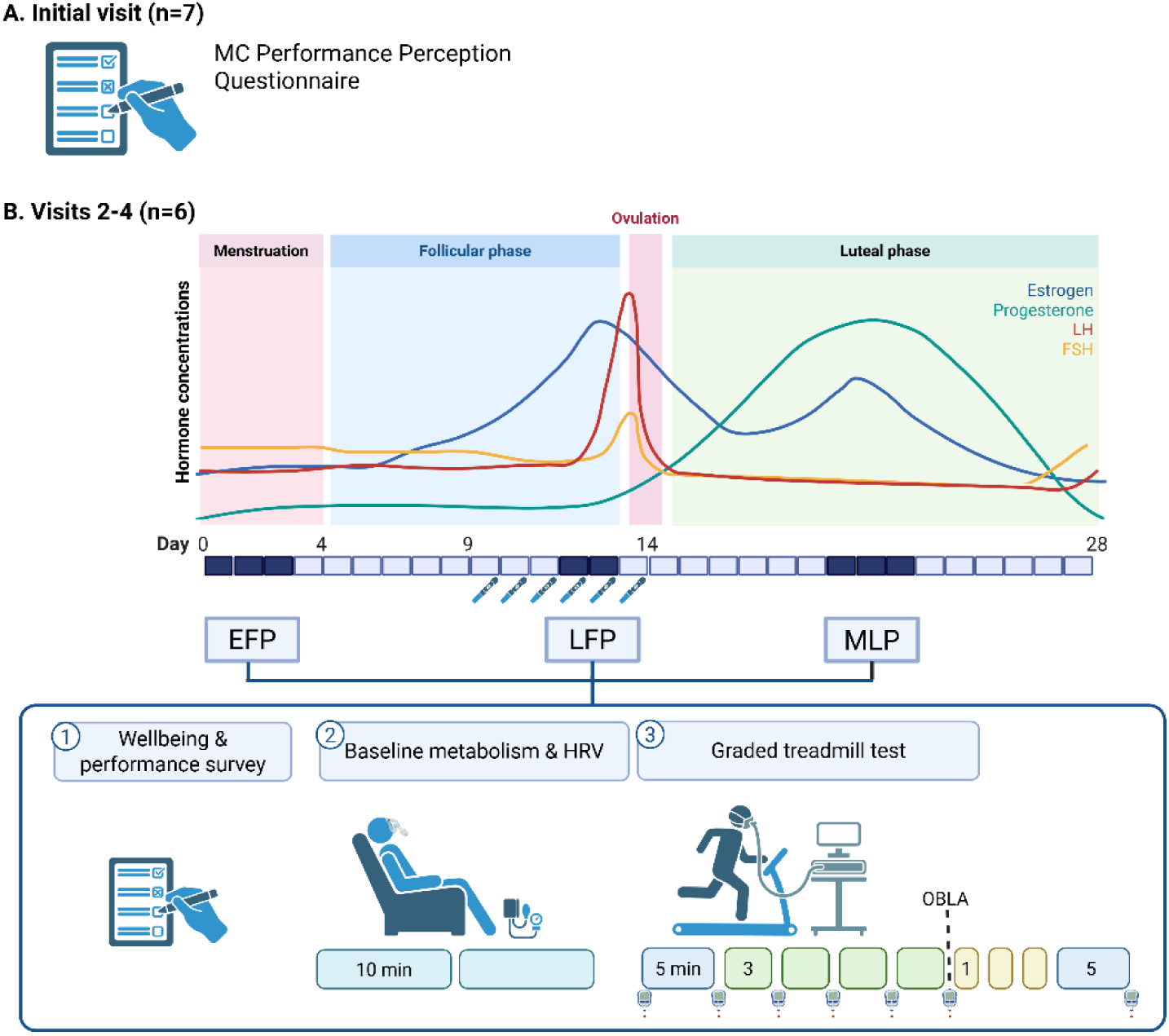
Schematic representation of the study design and protocol layout. A. Initial visit where participants completed a MC performance perception and past experiences questionnaire. B. Visits 2-4 completed during EFP: early follicular phase, LPF: late follicular phase, and MLP: mid-luteal phase. Ovulation test started 5 days before the expected ovulation day. On testing days, participants completed a wellbeing and performance perception questionnaire and conducted a resting test and graded test in which lactate was tested until reaching OBLA: Onset of Blood Lactate Accumulation.

### Menstrual-Cycle Phase Determination

MC phase was determined by combining menstrual calendar tracking, where both the participant and the researcher tracked the first day of the cycle and its duration, and urinary ovulation prediction kit (Premom ovulation test kit). Testing for luteinising hormone (LH) surges was carried out five days before anticipated ovulation. Results were interpreted using the Premom app, which identifies the LH surge approximately 24–36 hours before ovulation. Testing sessions were scheduled for days 1–3 of menses (EFP), one day before the LH peak (LFP), and 7–9 days post-ovulation (MLP). Participants recorded their test results in the Premom app and shared the data with the investigator.

### Testing Procedures

All testing sessions were conducted in the morning (06:00–09:00h) following an overnight fast and abstention from caffeine. One participant completed the graded test at 16:30 h due to scheduling constraints, but followed a minimum four-hour fast with no caffeine intake. Upon arrival, participants completed a short questionnaire (Appendix 1) assessing current menstrual symptoms, perceived performance, and general well-being. Resting blood pressure was recorded, followed by a 10-minute supine rest before RMR and HRV measurements.

#### - Resting Metabolic Rate and Heart Rate Variability

RMR was determined using indirect calorimetry via the VO₂ Master Pro gas-exchange system (VO₂ Master, Vernon, Canada). Participants remained still and awake in a reclined position for a continuous 10-minute recording period. HRV and resting heart rate were simultaneously recorded using a Polar H10 chest strap, with beat-to-beat R–R intervals analysed for autonomic variability.

#### - Graded Exercise Test

After the resting phase, participants completed a graded treadmill test to volitional exhaustion. Body mass and baseline blood lactate concentration (Lactate Pro 2, Arkray, Japan) were recorded before the test. The treadmill incline was set to 1% to mimic outside conditions^30^, and speed was hidden from participants to minimise pacing bias. Participants began with a 5-minute warm-up at approximately 55–60 % of their estimated VO₂max (based on recent 5 km performance). Thereafter, treadmill speed increased by 1 km·h⁻¹ every three minutes. Blood lactate was sampled at the end of each stage until the onset of blood lactate accumulation (OBLA, ∼4 mmol·L⁻¹). Once OBLA was reached, speed increments of 1 km·h⁻¹ per minute continued until exhaustion, Borg RPE 20, or voluntary termination. Perceived exertion (6–20 scale) was recorded after each stage. Heart rate was continuously monitored (Polar H10), and gas exchange was continuously measured via VO₂ Master Pro, with the average of the final minute per stage used for analysis.

### Statistical Analysis

Given the exploratory design no prior power calculation was performed. Descriptive statistics were used to explore trends across MC phases. Data distribution was assessed using Shapiro–Wilk tests and Q–Q plots. Parametric data were analysed using a one-way repeated-measures ANOVA with Tukey’s post-hoc comparisons, whereas non-parametric data were analysed with the Friedman test followed by Dunn’s multiple-comparison procedure. All analyses were performed using GraphPad Prism v10.5.0 (GraphPad Software, San Diego, USA).

## 3. Results

### 3.1 Participant Demographics and Characteristics

Ten female ultra-endurance athletes initially volunteered for the study. Seven completed the preliminary MC-related performance perception and previous experiences questionnaire, while six participants completed the physiological testing protocol and phase-specific questionnaires.

Descriptive demographic and training characteristics are summarised in Table 1.

**Table 1.** Participants characteristics (n=7)

| Variable | Mean ±SD |
| --- | --- |
| Age (years) | 37.14 ±6.6 |
| Body weight (kg) | 59.21 ±5.6 |
| Body height (cm) | 159.43 ±4.9 |
| VO<sub>2</sub>max (ml·kg<sup>-1</sup>·min<sup>-1</sup>) | 51.43 ±5.7 |
| Weekly training hours (h·wk<sup>-1</sup>) | 10.57 ±2.7 |
| Ultramarathon experience (years) | 2.57 ±1.1 |

Values are mean ± SD

### 3.2 Menstrual Cycle Impact on Performance Perception

Key MC characteristics are summarised in Table 2. The self-reported intensity of menstrual bleeding ranged from light (1) to heavy (3), with a mean score of 1.71 (SD ±0.76). Only one athlete reported substantial dysmenorrhea.

**Table 2.** MC characteristics (n=7)

| Variable | Mean ±SD |
| --- | --- |
| Age at menar | 12.71 ±1.9 |
| Length of the | 27.43 ±1.9 |
| Bleeding length | 4.21 ±0.8 |

Values are mean ± SD

Four out of seven female (57.1%) indicated that their MC affected performance, notably in the days before and during menstruation. The most common symptoms reported were fatigue, disturbed sleep, mood fluctuations, gastrointestinal issues, and reduced concentration. Several participants experienced these symptoms but did not perceive a direct decline in performance.

Two participants (28.6%) indicated that their MC had negatively affected race performance, and one reported withdrawing from a race due to menstrual-related symptoms. Qualitative responses indicated pacing difficulties, gastrointestinal discomfort, and frequent bathroom needs during events.

### 3.3 Physiological and Performance Outcomes

#### - Submaximal Performance

Submaximal efforts were evaluated at blood lactate concentrations of 2 mmol·L⁻¹ and 4 mmol·L⁻¹. There were no statistically significant differences in velocity, heart rate (HR), rate of perceived exertion (RPE), or running economy (RE) at either intensity across the different phases (Table 3).

**Table 3.** Performance characteristics during the MC phases (n=6)

| Variable | EFP |  |  | LFP |  |  | MLP |  |  | Effect of Phase | Mean within Participant CV |  |
| --- | --- | --- | --- | --- | --- | --- | --- | --- | --- | --- | --- | --- |
|  | Mean | ±SD | 95% CI | Mean | ±SD | 95% CI | Mean | ±SD | 95% CI | P |  | % |
| Body weight (Kg) | 59.58 | 6.24 | 53.03-66.13 | 59.23 | 6.62 | 52.29-66.18 | 59.30 | 6.62 | 52.35-66.25 | 0.52 |  | 0.93 |
| **Submaximal performance (2mmol/L Lactate)** |  |  |  |  |  |  |  |  |  |  |  |  |
| Velocity | 9.58 | 2.33 | 7.14-12.03 | 9.75 | 2.48 | 7.14-12.36 | 9.75 | 1.92 | 7.74-11.76 | 0.75 |  | 4.48 |
| HR | 145.75 | 8.34 | 137.0-154.5 | 147.50 | 11.43 | 135.5-159.5 | 148.56 | 8.27 | 139.9-157.2 | 0.74 |  | 3.94 |
| RPE | 12.17 | 2.14 | 9.92-14.41 | 11.83 | 2.32 | 9.40-14.26 | 12.50 | 1.97 | 10.43-14.57 | 0.60 |  | 9.28 |
| RE (ml·kg<sup>-1</sup>·km<sup>-1</sup>) | 236.09 | 39.65 | 180.4-290.3 | 221.59 | 25.26 | 192.1-258.1 | 231.69 | 35.35 | 196.2-281.5 | 0.39 |  | 7.39 |
| RF (breath per minute) | 41.88 | 6.45 | 35.11-48.66 | 39.56 | 9.17 | 29.93-49.19 | 43.23 | 6.39 | 36.53-49.93 | **0.045\*** |  | 6.26 |
| **Submaximal performance (4mmol/L Lactate)** |  |  |  |  |  |  |  |  |  |  |  |  |
| Velocity | 11.58 | 1.50 | 10.01-13.15 | 11.75 | 2.27 | 9.36-14.41 | 11.75 | 1.64 | 10.03-13.47 | 0.80 |  | 4.67 |
| HR | 162.44 | 6.89 | 155.2-169.7 | 169.58 | 3.85 | 165.5-173.6 | 166.71 | 6.38 | 160.0-173.4 | 0.08 |  | 3.06 |
| RPE | 16.00 | 0.63 | 15.34-16.66 | 15.83 | 1.94 | 13.80-17.87 | 17.00 | 1.10 | 15.85-18.15 | 0.23 |  | 7.18 |
| RE (ml·kg<sup>-1</sup>·km<sup>-1</sup>) | 229.85 | 29.87 | 198.5-261.2 | 227.21 | 24.72 | 201.3-253.2 | 229.81 | 26.57 | 201.9-257.7 | 0.96 |  | 7.64 |
| RF (breath per minute) | 47.00 | 5.79 | 40.93-53.08 | 46.24 | 5.92 | 40.02-52.45 | 50.03 | 7.96 | 41.67-58.38 | 0.25 |  | 5.69 |
| **Maximal Aerobic performance** |  |  |  |  |  |  |  |  |  |  |  |  |
| VO<sub>2peak</sub> (ml·kg<sup>-1</sup>·min<sup>-1</sup>) | 53.32 | 9.49 | 43.36-63.27 | 54.10 | 3.16 | 50.78-57.42 | 54.37 | 5.29 | 48.81-59.92 | 0.96 |  | 8.35 |
| Peak Velocity | 13.83 | 2.79 | 10.91-16.76 | 14.83 | 1.17 | 13.61-16.06 | 14.83 | 1.72 | 13.03-16.64 | 0.26 |  | 7.02 |
| TTE (min) | 22.61 | 4.20 | 18.20-27.02 | 24.08 | 2.48 | 21.48-26.68 | 24.75 | 2.52 | 22.10-27.40 | 0.99 |  | 5.75 |
| Peak HR | 175.33 | 3.83 | 171.3-179.4 | 181.83 | 9.54 | 171.8-191.8 | 180.33 | 8.52 | 171.4-189.3 | 0.53 |  | 2.65 |

Values are mean ± SD (95% Confidence Interval, CI). CV = coefficient of variation; HR, heart rate; RPE, rating of perceived exertion; RE = running economy; RF = respiratory frequency; VO<sub>2peak</sub> = peak oxygen uptake; HR = heart rate; TTE, time to exhaustion. Repeated-measures ANOVA or Friedman test applied as appropriate. \*p < 0.05.

A significant main effect of the MC phase was observed on RF at LT1 (*P* = 0.045), but pairwise comparison did not reveal a significant difference between phases. The largest effect was observed between LFP [39.6 ± 9.2 breath-per-minute (bpm)] and MLP (43.2 ± 6.6 bpm; *P* = 0.1). All other comparisons were not significant (*P*>0.05) (Table 3).

#### - Maximal Aerobic Performance

Maximal performance variables—including VO₂peak, peak treadmill velocity, time to exhaustion (TTE), and peak HR— were not different across the menstrual phases.

Slightly reduced performances were observed in the EFP due to two atypical cases: one participant ended the test early due to a migraine and light-headedness associated with menses, and another voluntarily discontinued upon reaching OBLA because of residual fatigue following a 9-hour mountain run completed 48 hours before the test.

Post-exercise blood lactate concentrations were not significantly different between phases; however, data from the EFP were excluded from this comparison due to the early test termination in these two cases (Table 3).

#### - Resting Physiological Characteristics

Resting variables – including resting HR, blood pressure, HRV, RMR or resting lactate – did not show any differences across the MC phases (Table 4).

**Table 4.** Health characteristics during the MC phases (n=6)

| Variable | EFP |  |  | LFP |  |  | MLP |  |  | Effect of Phase | Mean within Participant CV |
| --- | --- | --- | --- | --- | --- | --- | --- | --- | --- | --- | --- |
|  | Mean | ±SD | 95% CI | Mean | ±SD | 95% CI | Mean | ±SD | 95% CI | P | % |
| Resting HR | 54.17 | 6.05 | 47.82-60.51 | 58.00 | 8.76 | 48.80-67.20 | 57.33 | 5.47 | 51.60-63.07 | 0.39 | 8.48 |
| Sistolic Blood Pressure | 114.33 | 11.67 | 102.1-126.6 | 109.33 | 5.01 | 104.1-114.6 | 109.17 | 4.67 | 104.3-114.1 | 0.32 | 5.31 |
| Diastolic Blood Pressure | 70.00 | 3.35 | 66.49-73.51 | 71.00 | 4.24 | 66.55-75.45 | 67.83 | 4.31 | 63.31-72.36 | 0.26 | 3.23 |
| HRV | 64.48 | 5.72 | 56.95-67.60 | 67.40 | 6.30 | 55.51-78.29 | 64.08 | 6.89 | 53.47-70.03 | 0.25 | 6.53 |
| RMR (VO<sub>2</sub> (ml·kg<sup>-1</sup>·min<sup>-1</sup>))** | 3.58 | 0.28 | 3.29-3.88 | 3.75 | 0.56 | 3.17-4.34 | 3.98 | 0.69 | 3.26-4.70 | 0.38 | 10.66 |
| Resting Lactate | 1.47 | 0.20 | 1.26-1.67 | 1.58 | 0.48 | 1.08-2.09 | 1.43 | 0.25 | 1.17-1.70 | 0.58 | 12.05 |

Values are mean ± SD (95% Confidence Interval, CI). CV = coefficient of variation; HR, heart rate; HRV, heart rate variability; RMR, resting metabolic rate. Repeated-measures ANOVA or Friedman test applied as appropriate. No significant differences were observed between phases.

#### - Psychological and Perceptual Ratings

Phase-specific questionnaires on performance perception, energy, sleep quality, stress or anxiety, focus, and mood did not show significant differences across the menstrual phases (Table 5).

**Table 5.**
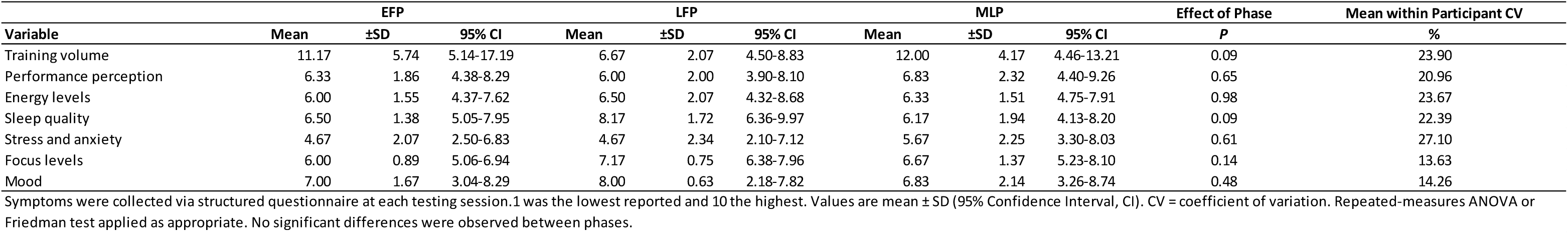
MC symptoms and psychological readiness (n=6)

| Variable | EFP |  |  | LFP |  |  | MLP |  |  | Effect of Phase | Mean within Participant CV |
| --- | --- | --- | --- | --- | --- | --- | --- | --- | --- | --- | --- |
|  | Mean | ±SD | 95% CI | Mean | ±SD | 95% CI | Mean | ±SD | 95% CI | P | % |
| Training volume | 11.17 | 5.74 | 5.14-17.19 | 6.67 | 2.07 | 4.50-8.83 | 12.00 | 4.17 | 4.46-13.21 | 0.09 | 23.90 |
| Performance perception | 6.33 | 1.86 | 4.38-8.29 | 6.00 | 2.00 | 3.90-8.10 | 6.83 | 2.32 | 4.40-9.26 | 0.65 | 20.96 |
| Energy levels | 6.00 | 1.55 | 4.37-7.62 | 6.50 | 2.07 | 4.32-8.68 | 6.33 | 1.51 | 4.75-7.91 | 0.98 | 23.67 |
| Sleep quality | 6.50 | 1.38 | 5.05-7.95 | 8.17 | 1.72 | 6.36-9.97 | 6.17 | 1.94 | 4.13-8.20 | 0.09 | 22.39 |
| Stress and anxiety | 4.67 | 2.07 | 2.50-6.83 | 4.67 | 2.34 | 2.10-7.12 | 5.67 | 2.25 | 3.30-8.03 | 0.61 | 27.10 |
| Focus levels | 6.00 | 0.89 | 5.06-6.94 | 7.17 | 0.75 | 6.38-7.96 | 6.67 | 1.37 | 5.23-8.10 | 0.14 | 13.63 |
| Mood | 7.00 | 1.67 | 3.04-8.29 | 8.00 | 0.63 | 2.18-7.82 | 6.83 | 2.14 | 3.26-8.74 | 0.48 | 14.26 |

Symptoms were collected via structured questionnaire at each testing session. 1 was the lowest reported and 10 the highest. Values are mean ± SD (95% Confidence Interval, CI). CV = coefficient of variation. Repeated-measures ANOVA or Friedman test applied as appropriate. No significant differences were observed between phases.

## 4. Discussion

This pilot study investigated the influence of MC phases (EFP, LFP, and MLP) on physiological and perceptual responses in trained female ultra-endurance athletes. To our knowledge, this is the first study to investigate the variation across the MC in submaximal, maximal performance variables, autonomic function, metabolism and self-perceived outcomes in this population. Understanding potential hormonal effects in ultra-endurance contexts is increasingly relevant as female participation continues to rise.

57.1% of the participants initially reported that the MC affected their performance, particularly before and during menstruation, with common symptoms including fatigue, disturbed sleep, mood changes, gastrointestinal distress, and reduced focus. These symptoms were not always associated with worse performance perception. This is in agreement with previous studies on elite athletes where perceptual disruption is common despite no change in performance ^29,31–33^. This aligns with data from track and field populations that also reported worse perceived performance in the late luteal and early follicular phase^29^.

No statistically significant differences were observed in submaximal (HR response, RE, RPE or submaximal velocities) or maximal (VO₂peak, HRmax, time-to-exhaustion or peak velocity) variables in the different MC-phases. These results align with prior research showing minimal physiological fluctuations across the cycle in trained female athletes^34^, and extend the findings of the FENDURA project^27^ to a novel ultra-endurance cohort.

A small but notable phase effect was detected for respiratory frequency (RF) at LT1, although post-hoc comparisons did not reach significance, with the largest difference seen between the LFP and MLP. These findings could suggest a potential trend towards greater ventilatory efficiency under higher oestrogen and lower progesterone concentrations, consistent with earlier reports of improved running economy in the follicular phase^35^. Given the prolonged, submaximal nature of ultra-endurance exercise, even minor improvements in ventilatory or metabolic efficiency could have practical implications for fatigue resistance. However, the small sample size precludes firm conclusions, and this may be a focus area for future research.

Endogenous oestrogen is known to enhance nitric oxide–mediated vasodilation and mitochondrial oxidative capacity, promoting lipid oxidation and glycogen sparing during submaximal exercise^6,14–17,36^. In contrast, the lower progesterone concentrations during the LFP reduce the thermogenic and ventilatory drive typically seen in the luteal phase^5^, potentially decreasing perceived exertion. Such mechanisms may underlie the subtle improvements in ventilatory or metabolic efficiency observed in our study and in other work reporting similar trends^3^.

Similar findings were reported by Rael *et al.* (2021)^3^, observing a modest but significant phase-related difference in ventilation, relative VO₂, and energy expenditure during low-intensity recovery exercise, with improved efficiency during the LFP compared with both the EFP and MLP^3^. In another study, Goldsmith and Glaister (2021)^35^ reported poorer running economy during the luteal phase, although their late-follicular window was defined as the day before predicted ovulation without hormonal confirmation, which may have underestimated the true high-oestrogen period. In contrast, the FENDURA Project did not find meaningful differences across the MC, despite more rigorous hormonal validation^37^. Although, it is worth noting that their “high-oestrogen” phase was identified as the day after a positive LH test^37^, by which time circulating oestrogen levels are already declining^38,39^. These methodological discrepancies in phase definition and verification could partly explain the inconsistency across studies.

Resting HR, HRV, RMR and lactate concentration did not differ across the phases. In nonathletic populations, a shift towards sympathetic dominance is observed linked to changes in perceptual states^40–42^. The lack of difference in our population could reflect a high degree of cardiovascular adaptation typical seen in ultra-endurance athletes. Chronic endurance training enhances vagal tone and decreases day-to-day HRV variability^43^, which may attenuate or mask the influence of cyclical hormonal variations on resting autonomic function. This autonomic stability could also explain the lack of significant phase-related variation in subjective wellbeing parameters—including perceived energy, sleep quality, stress, anxiety, and performance perception. Although minor improvements were noted during the late follicular phase, these did not reach statistical significance, most likely due to inter-individual variability and the small sample size. Collectively, these findings suggest that the high physiological conditioning typical of ultra-endurance athletes may buffer any subtle menstrual-phase influences on both autonomic and perceptual outcomes.

Accurate MC phase determination remains a major challenge in exercise endocrinology. While serum hormone verification is considered the gold standard^26^, practical constraints often preclude its use in small field-based studies. The combined calendar and urinary LH approach adopted here improves on calendar-only methods but cannot identify anovulatory or luteal-deficient cycles. Variability in phase definitions across studies may further obscure transient hormonal effects. Continued refinement of verification protocols is therefore essential to enhance comparability and interpretation across studies.

## 5. Limitations and Future Directions

The small sample size (n = 6/7) and heterogeneity in age and training load limit generalisability and statistical power. Allowing athletes to maintain habitual training schedules enhanced ecological validity but introduced uncontrolled variation. The use of a graded treadmill protocol may also be less reflective of the prolonged submaximal intensities typical of ultramarathon performance. Future research should aim for larger longitudinal cohorts including at least 3 consecutive cycles, hormonal confirmation through serum analysis, and event-specific performance tests, possibly under field conditions.

## 6. Conclusions

This study found no significant effects of menstrual-phase on physiological performance in trained female ultra-endurance athletes. Nonetheless, over half of the participants perceived performance decrements around menstruation, underscoring the relevance of perceptual monitoring; however, this perception was not observed when phase-specific questionnaires were answered. The findings highlight high inter-individual variability and support the inclusion of personalised MC tracking within endurance-training programmes. Continued research using validated phase identification and combined physiological-perceptual outcomes is needed to clarify how hormonal fluctuations interact with adaptation and performance in ultra-endurance athletes.

## Data Availability

All data produced in the present study are available upon reasonable request to the authors

## Acknowledgments

The authors would like to thank Aranzazu Peñaranda López for granting access to the performance testing facilities at the Manual Therapy Clinic and the VO2Master team for their valuable technical support throughout the project.

## Additional information

## Illustrations

Figures created with BioRender.com. License number: *XH28XVFCR1*

## References

1 Ose, B. M. et al. Where Are All the Female Participants in Sports and Exercise Medicine Research? A Decade Later. Am J Sports Med 53, 2022–2028 (2025). 10.1177/03635465241278350

2 Benton, M. J., Hutchins, A. M. & Dawes, J. J. Effect of menstrual cycle on resting metabolism: A systematic review and meta-analysis. PLoS One 15, e0236025 (2020). 10.1371/journal.pone.0236025

3 Rael, B. et al. Menstrual Cycle Phases Influence on Cardiorespiratory Response to Exercise in Endurance-Trained Females. International Journal of Environmental Research and Public Health 18, 860 (2021).

4 Lavanya, R., Sureshbalaji, R. A. & Prem Kumar, S. Cardiac Efficiency and Work Performance Variations Across Menstrual Cycle Phases: A Bicycle Ergometric Study in Young Women. Cureus 17, e78216 (2025). 10.7759/cureus.78216

5 Janse, D. E. J. X. A., Thompson, M. W., Chuter, V. H., Silk, L. N. & Thom, J. M. Exercise performance over the menstrual cycle in temperate and hot, humid conditions. Med Sci Sports Exerc 44, 2190–2198 (2012). 10.1249/MSS.0b013e3182656f13

6 Hackney, A. C. Menstrual Cycle Hormonal Changes and Energy Substrate Metabolism in Exercising Women: A Perspective. Int J Environ Res Public Health 18 (2021). 10.3390/ijerph181910024

7 Fort-Vanmeerhaeghe, A. et al. Injury Risk and Overall Well-Being During the Menstrual Cycle in Elite Adolescent Team Sports Athletes. Healthcare (Basel) 13 (2025). 10.3390/healthcare13101154

8 Costa, R. J. S., Knechtle, B., Tarnopolsky, M. & Hoffman, M. D. Nutrition for Ultramarathon Running: Trail, Track, and Road. Int J Sport Nutr Exerc Metab 29, 130–140 (2019). 10.1123/ijsnem.2018-0255

9 Knechtle, B. Ultramarathon Runners: Nature or Nurture? International journal of sports physiology and performance 7, 310–312 (2012). 10.5167/uzh-67287

10 Pappa, D., et al. Effect of an ultra-distance foot race on the hypothalamic-pituitary testicular adrenocortical axes insulin growth factor levels: distinct patterns of suppression, stimulation and recovery. Annals of Research Hospitals 3 (2019).

11 Le Goff, C., Gergelé, L., Seidel, L., Cavalier, E. & Kaux, J.-F. Mountain Ultra-Marathon (UTMB) Impact on Usual and Emerging Cardiac Biomarkers. Frontiers in Cardiovascular Medicine **Volume** 9 - 2022 (2022). 10.3389/fcvm.2022.856223

12 UTMB®. UTMB World Series 2024: season in figures, <https://utmb.world/news/announcement-key-figures-2024> (2024).

13 Carmichael, M. A., Thomson, R. L., Moran, L. J. & Wycherley, T. P. The Impact of Menstrual Cycle Phase on Athletes’ Performance: A Narrative Review. Int J Environ Res Public Health 18 (2021). 10.3390/ijerph18041667

14 Moreau, K. L., Stauffer, B. L., Kohrt, W. M. & Seals, D. R. Essential Role of Estrogen for Improvements in Vascular Endothelial Function With Endurance Exercise in Postmenopausal Women. The Journal of Clinical Endocrinology & Metabolism 98, 4507–4515 (2013). 10.1210/jc.2013-2183

15 SenthilKumar, G., Katunaric, B., Bordas-Murphy, H., Sarvaideo, J. & Freed, J. K. Estrogen and the Vascular Endothelium: The Unanswered Questions. Endocrinology 164 (2023). 10.1210/endocr/bqad079

16 Duckles, S. P. & Miller, V. M. Hormonal modulation of endothelial NO production. Pflugers Arch 459, 841–851 (2010). 10.1007/s00424-010-0797-1

17 Pellegrino, A., Tiidus, P. M. & Vandenboom, R. Mechanisms of Estrogen Influence on Skeletal Muscle: Mass, Regeneration, and Mitochondrial Function. Sports Med 52, 2853–2869 (2022). 10.1007/s40279-022-01733-9

18 Oosthuyse, T. & Bosch, A. N. The effect of the menstrual cycle on exercise metabolism: implications for exercise performance in eumenorrhoeic women. Sports Med 40, 207–227 (2010). 10.2165/11317090-000000000-00000

19 Löfberg, I. E. et al. Peak Fat Oxidation during Submaximal Exercise Remains Consistent across Menstrual Cycle and Combined Oral Contraceptive Phases. Med Sci Sports Exerc 57, 1383–1394 (2025). 10.1249/mss.0000000000003676

20 D’Souza, A. C. et al. Menstrual cycle hormones and oral contraceptives: a multimethod systems physiology-based review of their impact on key aspects of female physiology. Journal of Applied Physiology 135, 1284–1299 (2023). 10.1152/japplphysiol.00346.2023

21 Kumagai, H. et al. Genetic polymorphisms in CYP19A1 and ESR1 are associated with serum CK activity after prolonged running in men. Journal of Applied Physiology 132, 966–973 (2022). 10.1152/japplphysiol.00374.2021

22 Dombovy, M. L., Bonekat, H. W., Williams, T. J. & Staats, B. A. Exercise performance and ventilatory response in the menstrual cycle. Medicine & Science in Sports & Exercise 19, 111–117 (1987).

23 Jurkowski, J. E., Jones, N. L., Toews, C. J. & Sutton, J. R. Effects of menstrual cycle on blood lactate, O2 delivery, and performance during exercise. Journal of Applied Physiology 51, 1493–1499 (1981). 10.1152/jappl.1981.51.6.1493

24 Greenhall, M., Taipale, R. S., Ihalainen, J. K. & Hackney, A. C. Influence of the Menstrual Cycle Phase on Marathon Performance in Recreational Runners. Int J Sports Physiol Perform 16, 601–604 (2021). 10.1123/ijspp.2020-0238

25 McNulty, K. L. et al. The Effects of Menstrual Cycle Phase on Exercise Performance in Eumenorrheic Women: A Systematic Review and Meta-Analysis. Sports Med 50, 1813–1827 (2020). 10.1007/s40279-020-01319-3

26 Schaumberg, M. A., Jenkins, D. G., Janse de Jonge, X. A. K., Emmerton, L. M. & Skinner, T. L. Three-step method for menstrual and oral contraceptive cycle verification. Journal of Science and Medicine in Sport 20, 965–969 (2017). 10.1016/j.jsams.2016.08.013

27 Taylor, M. Y. et al. Menstrual Cycle Phase Has No Influence on Performance-Determining Variables in Endurance-Trained Athletes: The FENDURA Project. Med Sci Sports Exerc 56, 1595–1605 (2024). 10.1249/mss.0000000000003447

28 Brown, N., Knight, C. J. & Forrest Née Whyte, L. J. Elite female athletes’ experiences and perceptions of the menstrual cycle on training and sport performance. Scand J Med Sci Sports 31, 52–69 (2021). 10.1111/sms.13818

29 Jones, B. P. et al. Menstrual cycles and the impact upon performance in elite British track and field athletes: a longitudinal study. Frontiers in Sports and Active Living 6 (2024). 10.3389/fspor.2024.1296189

30 Jones, A. M. & Doust, J. H. A 1% treadmill grade most accurately reflects the energetic cost of outdoor running. J Sports Sci 14, 321–327 (1996). 10.1080/02640419608727717

31 Bruinvels, G. et al. Prevalence and frequency of menstrual cycle symptoms are associated with availability to train and compete: a study of 6812 exercising women recruited using the Strava exercise app. Br J Sports Med 55, 438–443 (2021). 10.1136/bjsports-2020-102792

32 Oester, C. et al. Inconsistencies in the perceived impact of the menstrual cycle on sport performance and in the prevalence of menstrual cycle symptoms: A scoping review of the literature. Journal of Science and Medicine in Sport 27, 373–384 (2024). 10.1016/j.jsams.2024.02.012

33 Prado, R. C. R., Silveira, R., Kilpatrick, M. W., Pires, F. O. & Asano, R. Y. The effect of menstrual cycle and exercise intensity on psychological and physiological responses in healthy eumenorrheic women. Physiology & Behavior 232, 113290 (2021). 10.1016/j.physbeh.2020.113290

34 Meignié, A. et al. The Effects of Menstrual Cycle Phase on Elite Athlete Performance: A Critical and Systematic Review. Frontiers in Physiology **Volume** 12 - 2021 (2021). 10.3389/fphys.2021.654585

35 Goldsmith, E. & Glaister, M. The effect of the menstrual cycle on running economy. J Sports Med Phys Fitness 60, 610–617 (2020). 10.23736/s0022-4707.20.10229-9

36 Yoh, K., Ikeda, K., Horie, K. & Inoue, S. Roles of Estrogen, Estrogen Receptors, and Estrogen-Related Receptors in Skeletal Muscle: Regulation of Mitochondrial Function. Int J Mol Sci 24 (2023). 10.3390/ijms24031853

37 Docter, H. et al. Running Economy After a Low- and High-Intensity Training Session in Naturally Menstruating Endurance-Trained Female Athletes: The FENDURA Project. Scandinavian Journal of Medicine & Science in Sports 35, e70050 (2025). 10.1111/sms.70050

38 Maman, E., Adashi, E. Y., Baum, M. & Hourvitz, A. Prediction of ovulation: new insight into an old challenge. Scientific Reports 13, 20003 (2023). 10.1038/s41598-023-47241-2

39 Stricker, R. et al. Establishment of detailed reference values for luteinizing hormone, follicle stimulating hormone, estradiol, and progesterone during different phases of the menstrual cycle on the Abbott ARCHITECT analyzer. Clin Chem Lab Med 44, 883–887 (2006). 10.1515/cclm.2006.160

40 de Zambotti, M., Nicholas, C. L., Colrain, I. M., Trinder, J. A. & Baker, F. C. Autonomic regulation across phases of the menstrual cycle and sleep stages in women with premenstrual syndrome and healthy controls. Psychoneuroendocrinology 38, 2618–2627 (2013). 10.1016/j.psyneuen.2013.06.005

41 Schmalenberger, K. M. et al. Menstrual Cycle Changes in Vagally-Mediated Heart Rate Variability are Associated with Progesterone: Evidence from Two Within-Person Studies. J Clin Med 9 (2020). 10.3390/jcm9030617

42 Schmalenberger, K. M. et al. A Systematic Review and Meta-Analysis of Within-Person Changes in Cardiac Vagal Activity across the Menstrual Cycle: Implications for Female Health and Future Studies. J Clin Med 8 (2019). 10.3390/jcm8111946

43 Plews, D. J., Laursen, P. B., Stanley, J., Kilding, A. E. & Buchheit, M. Training Adaptation and Heart Rate Variability in Elite Endurance Athletes: Opening the Door to Effective Monitoring. Sports Medicine 43, 773–781 (2013). 10.1007/s40279-013-0071-8

