## Supplementary Data for "Physiological and Perceptual Responses Across Menstrual Cycle Phases in Female Ultra-Endurance Athletes: A Pilot Study"

### QUESTIONNAIRE ON SYMPTOMS AND PERFORMANCE PERCEPTION

**Impact of Menstrual Cycle Hormonal Fluctuations on Performance in Female Ultra-Endurance Athletes**

**Participant number:**

**Questions to fill on Visit 1: Date -**

1) Height:
2) Weight:

3) What is your age?

4) What is your ethnicity?

5) Are you currently using any contraception?
   Yes
   No

6) Age at menarche?

7) Are your menstrual cycles regular?
   Yes
   No, *please provide details*

8) How long is your average menstrual cycle?

9) Do you have a history of amenorrhoea (absence of menstruation)?
   Yes, *please provide details*
   No

10) Have you ever been diagnosed with any metabolic, cardiovascular, hormonal disorders or any condition affecting the reproductive system?
Yes
No

11) How long have you been competing in ultramarathons?

12) What's your current training volume?

13) Do you know your VO2 max? If so, please include it and indicate whether it's a watch metric or a lab result.

14) On average, how long does your period last (bleeding days)?

15) How would you describe your menstrual bleeding?
- ☐ Light
- ☐ Moderate
- ☐ Heavy

16) Do you experience significant menstrual pain?
Yes
No

17) Do you bleed between your periods?
Yes
No

18) Do you suffer from any of the following symptoms related to your menstrual cycle?
- ☐ Bloating

- Fatigue
- Gastrointestinal issues
- Lower back pain
- Abdominal cramps
- Sleep disturbance
- Headaches or migraines
- Weight fluctuations
- Muscle and/or joint pain
- Lack of focus
- Mood swings, *please indicate*
- Other, *please indicate*

19) Does your menstrual cycle affect your performance?

Yes

No

20) If so, when does this affect you?

- Before bleeding
- During bleeding
- Other time, *please indicate*

21) Has your menstrual cycle ever affected your performance in a competition?

Yes

No

*If yes, can you develop?*

22) Has your menstrual cycle ever caused you to withdraw from a competition?

Yes

No

*If yes, can you develop?*

23) Have you ever felt you get injuries or niggles in certain stages of your menstrual cycle?

Yes

No

*If yes, can you develop?*

**Questions to fill on Visit 2: Date -**

1) Total training volume of this last week:

2) What phase of your menstrual cycle are you currently in?
    - ☐ Menstruation
    - ☐ Follicular phase
    - ☐ Luteal phase

3) How would you rate your performance on a scale of 1-10 during the five days before this appointment

4) How would you rate your energy levels on a scale of 1-10, five days before this appointment?

5) Did you experience any niggles five days before this appointment?

6) Did you experience a bigger appetitive five days before this appointment?

7) Rate the quality of your sleep on a scale of 1-10, five days before this appointment:

8) Rate your stress/anxiety levels on a scale of 1-10, five days before this appointment:

9) Rate your focus levels on a scale of 1-10, five days before this appointment:

10) Rate mood on a scale of 1-10 (1 being very sad and 10 being very happy), five days before this appointment

**Questions to fill on Visit 3: Date -**

1) Total training volume of this last week:

2) What phase of your menstrual cycle are you currently in?
   - ☐ Menstruation
   - ☐ Follicular phase
   - ☐ Luteal phase

3) How would you rate your performance on a scale of 1-10 during the five days before this appointment

4) How would you rate your energy levels on a scale of 1-10, five days before this appointment?

5) Did you experience any niggles five days before this appointment?

6) Did you experience a bigger appetitive five days before this appointment?

7) Rate the quality of your sleep on a scale of 1-10, five days before this appointment:

8) Rate your stress/anxiety levels on a scale of 1-10, five days before this appointment:

9) Rate your focus levels on a scale of 1-10, five days before this appointment:

10) Rate mood on a scale of 1-10 (1 being very sad and 10 being very happy), five days before this appointment

**Questions to fill on Visit 4: Date -**

1) Total training volume of this last week:

2) What phase of your menstrual cycle are you currently in?
   - ☐ Menstruation
   - ☐ Follicular phase
   - ☐ Luteal phase

3) How would you rate your performance on a scale of 1-10 during the five days before this appointment

4) How would you rate your energy levels on a scale of 1-10, five days before this appointment?

5) Did you experience any niggles five days before this appointment?

6) Did you experience a bigger appetitive five days before this appointment?

7) Rate the quality of your sleep on a scale of 1-10, five days before this appointment:

8) Rate your stress/anxiety levels on a scale of 1-10, five days before this appointment:

9) Rate your focus levels on a scale of 1-10, five days before this appointment:

10) Rate mood on a scale of 1-10 (1 being very sad and 10 being very happy), five days before this appointment

Questionnaire adapted from Jones, B. P. *et al* (2024)[29]
